# Age-Related Modifications of Muscle Synergies during Daily-Living Tasks: A Scoping Review

**DOI:** 10.1101/2023.06.16.23291230

**Authors:** Cloé Dussault-Picard, Sara Havashinezhadian, Nicolas A Turpin, Florent Moissenet, Katia Turcot, Yosra Cherni

## Abstract

**Purpose:** Aging is associated with changes in neuromuscular control that can lead to difficulties in performing daily-living tasks. Based on electromyography, muscle synergy analysis provides a valuable tool for assessing neuromuscular control strategies. However, the age-related changes of muscle synergies during daily living tasks are scattered throughout the literature. This review aimed to synthesize the existing literature on muscle synergies in elderly people during daily-living tasks and examine how they differ from those exhibited by young adults.

**Methods:** A search was conducted across the Medline, CINHAL, and Web of Science databases. Studies were included if they focused on muscle synergies in elderly people during walking, sit-to-stand or stair ascent, and if muscle synergies were obtained by a matrix factorization algorithm.

**Results:** The research strategy identified 4849 studies, of which 17 studies were included after the screening process. The muscle synergies of 295 elderly people and 182 young adults were reported. Results suggest that: 1) elderly people and young adults retain similar muscle synergies’ number, 2) elderly people have higher muscles contribution during walking, and 3) an increased inter and intra-subject variability during specific tasks (i.e., walking and stair ascent, respectively) was reported in elderly people compared to young adults.

**Conclusion:** This review provides a comprehensive understanding of age-related changes in neuromuscular control during daily-living tasks. Our findings suggested that although the number of synergies remains similar, metrics such as spatial and temporal structures of synergies are more suitable to identify neuromuscular control deficits between young adults and elderly people.

## 1. Introduction

Aging is associated with changes in neuromuscular control (Schmitz et al., 2009), which refers to the coordinated interaction between the nervous system and the muscles. These alterations can have a major impact on mobility capacities (Brown and Flood, 2013). The combination of motor and cognitive disorders is at the origin of an accelerated loss of independence and autonomy (Bimou et al., 2021; Sobral et al., 2018). Additionally, the aging process is often accompanied by degeneration of nerve and muscle tissues. As a result, the performance of daily tasks (e.g., walking, sit-to-stand and stair ascent) becomes increasingly challenging for elderly people (EP). Indeed, performing these tasks are considered complex considering multi-level joints coordination and the need to coordinate different agonist and antagonist muscles. However, daily-living tasks are necessary skills to maintain independence and autonomy (Merrilees, 2014).

Muscle synergy analysis is recognized as a useful tool to assess neuromuscular control strategies or to quantify functional deficits in pathologies (Turpin et al., 2021). A muscle synergy analysis gives an insight into the temporal and spatial structure of the muscle’s coordination from the recorded muscle activity. So far, the most appropriate method to retrieve muscle synergies is by extracting muscle activity from electromyography (EMG) signals with the non-negative matrix factorization algorithm (Rabbi et al., 2020, Turpin et al., 2021). This method has been commonly used for assessing muscle synergies during daily-living tasks in various populations with muscle coordination impairments (Cherni et al., 2021; Turpin et al., 2021). Indeed, neuromuscular control impairments during functional tasks assessed by muscle synergies may be relevant to develop training modalities that are specific to the deficient synergies, especially in populations that suffer from neuromuscular control degradation such as EP.

The literature suggests that aging impacts how spinal circuits integrate peripheral afference and descending inputs, resulting in a change in final motor output (e.g., muscles synergies) in EP. For example, Baggen et al., (2020), found that neuromuscular control complexities and structures were affected by age during step ascent at different heights. Indeed, age was correlated with higher synergy complexity, and authors reported higher synergies similarity across step heights in the older compared to the young adults (YA) (Baggen et al., 2020). On the other hand, Monaco et al. (Monaco et al., 2010) reported that the gross structures of muscle synergies and their temporal activations were similar between YA and EP during locomotion, while Kubota et al. (2021) reported a decreased synergy complexity in EP, compared to YA. The contradictory results of the above studies concerning the effect of age on synergies complexity show that it remains unclear whether the between-group neuromuscular differences are attributed to changes in specific muscle synergies, their temporal activities, or both. Indeed, the studies that have investigated the relationship between aging and changes in synergies during daily living tasks are scattered, and a scoping of the literature is necessary to provide a better understanding of the effect of aging on muscle synergies during common daily living tasks (i.e., walking, sit-to stand, stair ascent). This would help to guide interventions in aging populations and ultimately, lighten the decline in self-mobility and autonomy.

This scoping review aims to give an overview of the existing studies investigating lower limb muscle synergies in EP during daily living tasks such as walking, sit-to-stand task and stair ascent. The primary aim is to examine how muscle synergies in EP differ from those exhibited by YA during walking, stair ascent, and sit-to-stand tasks by investigating the quantification and structure of synergies, and the variability of synergies between and within EP.

## 2. Materials and Methods

### 2.1. Data Source and Literature Source

A science librarian was consulted for the initial development of the search protocol. Studies were identified by searching Medline, CINHAL and Web of Science from inception to October 2022. The search strategy was based on three main concepts: “muscle synergy,” “elder, and “daily living tasks”. More details concerning search strategy and the key words used are reported in **Appendix** as a supplementary material. The current review follows the Systematic reviews and Meta-Analyses extension for Scoping Reviews (PRISMA-ScR) checklist (Tricco et al., 2018) and was registered on the OSF platform (ID: osf.io/e3bzv).

### 2.2. Eligibility Criteria

The included studies met the following inclusion criteria: (1) performing on a group of adults with a mean age of 60 years or older (as defined by the United Nations); without a history of major physical or psychiatric condition likely to affect gait, and in case of a mixed population: the majority of the investigated population older than 60; (2) focused on muscle synergies of the lower limb during walking, sit-to-stand task or stair ascent; (3) based on non-negative matrix factorization (NNMF) synergy extraction method and; (4) study published in French or English. Studies were excluded if they: (1) was performed on a population other than elder; (2) focused on muscle synergies of the upper limb; and (3) was not original research, such as letters to editor, conference abstracts and commentaries.

### 2.3. Studies screening

Titles and abstracts of the identified studies were screened independently by two of the authors (YC and SH) to identify those that potentially met inclusion criteria. A full review of those studies was then performed independently by the same authors. In the case of any unresolvable disagreement related to the studies eligibility, a third author (FM) performed the screening to reach consensus.

### 2.4. Methodological quality and risk of bias

Two authors independently (YC and SH) rated the overall quality of each included study, using the modified version of the Downs and Black checklist (Connor Gorber et al., 2007; Downs and Black, 1998). Out of 27 Items, fourteen Items were identified as relevant by the authors which allows to evaluate overall reporting bias (items 1, 2, 3, 4, 5, 6, 7, 10), external validity (items 11 and 12), internal validity bias (items 15, 16, 18, 20), internal validity confounding (items 21, 22, 25), and power (item 27) of the included studies. The maximum total consists of 19 points per study. Each study was assigned a score of “high” (≥75%), “moderate” (60–74%), “low” (≤60%) (Desmyttere et al., 2018). For the assessment procedures, a calibration meeting was initially performed with five studies, to ensure a clear understanding of each criterion and thus standardization and reliability of assessments. A second meeting was held to discuss the criteria for each study included, until a consensus was reached for a score. In the case of any unresolvable disagreement, a third author (FM) performed the assessment to reach consensus.

### 2.5. Data charting process

Data including study design, quality assessment, subject characteristics (age, sex), study methods (number of cycles analyzed, number and name of muscles recorded, EMG pre-processing methods), and synergy outcomes (muscle synergies in EP, and differences with YA), was extracted by one author (CDP), and validated by a second author (YC). Descriptive and numerical analyses were used to summarize the literature for each functional task (i.e., walking, sit-to stand, stair ascent). The main outcome measures discussed in this review were: (1) quantification of muscle synergies such as total number of synergies, the spatial (i.e., muscle weighting) and temporal (i.e., relative temporal activation) structure of muscle synergies, and the variability accounted for (VAF). The spatial and temporal structure of muscle synergies were reported as mentioned in the original article by the authors or extracted from the article graphics. The spatial and temporal structure of muscle synergies were reported as mentioned in the original article by the authors or extracted from the article graphics. If extracted from graphics, muscles with the highest weight and the most significant timing were reported to define the spatial and temporal structure, respectively. The VAF was defined by the uncentered Pearson correlation coefficient between weight x coefficient, and the EMG amplitude time series (Torres-Oviedo et al., 2006). Effect sizes were reported for each significant synergy difference between group (EP vs YA). If the original study does not provide the effect size, it was calculated from mean and standard deviation data. The authors were contacted if mean and standard deviation were not available. Cohen’s d effect size (d) or Glass’s delta effect size (△) was calculated if the study used parametric tests or non-parametric, respectively (Cohen, 1977; Ialongo, 2016). The findings related to the study aims and the implication for future research were then discussed.

## 3. Results

### 3.1. Search results

The initial search led to 8 963 studies. After removing duplicates, study titles and abstracts were screened by two reviewers to assess the eligibility of 4 849 studies. Then, 280 studies were determined by consensus and qualified for the full-text reading stage. This last stage resulted in the identification of 17 studies as eligible in this review. The flowchart of the selection process is charted in **Figure 1**.

**Figure 1.**
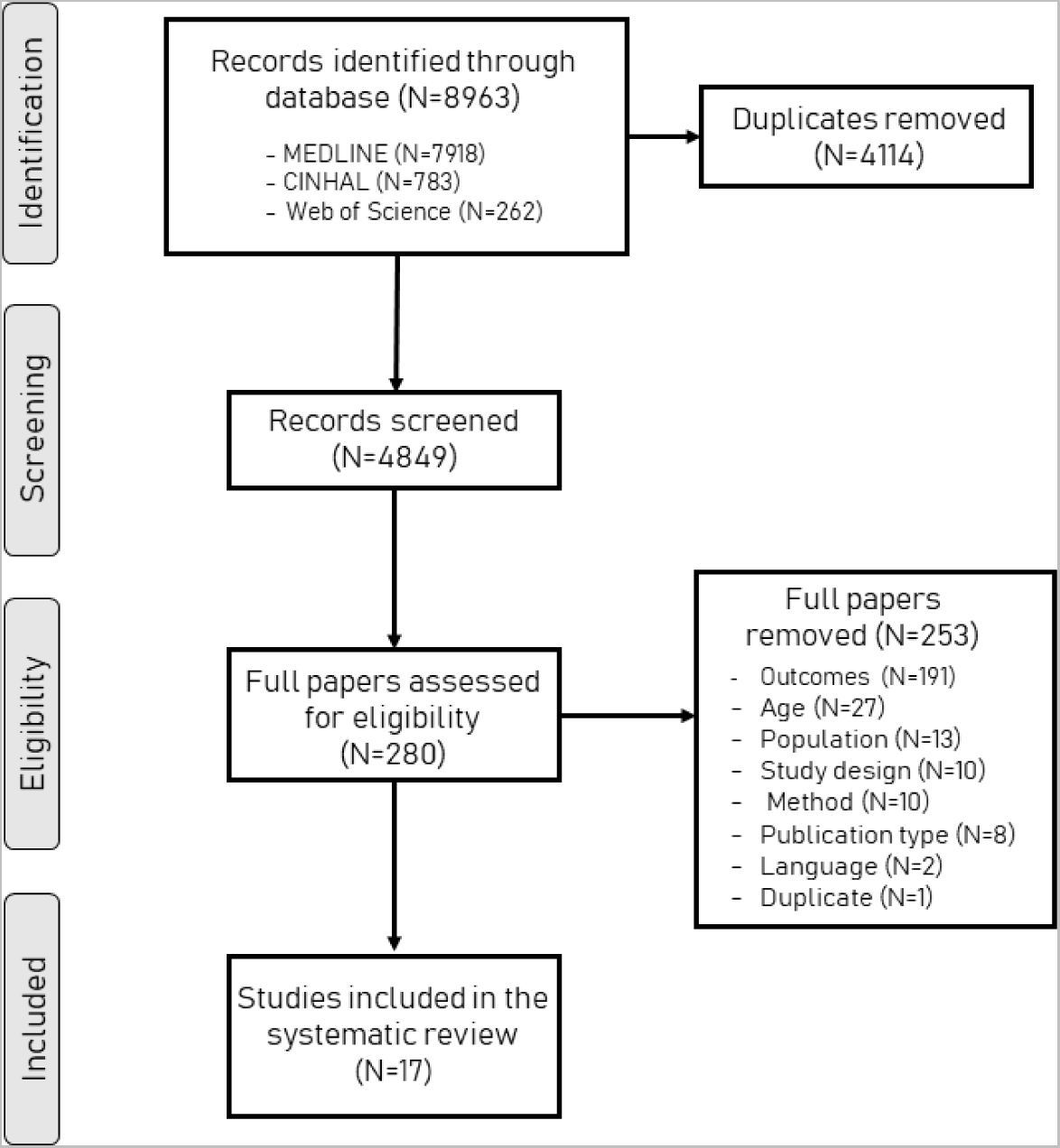
The scoping review flow diagram.

### 3.2. Risk of Bias

The median score of the modified Quality Index for the included studies was 72% (range from 44 to 89%) indicating a high quality (Table 1). The majority of studies were of high (Alizadehsaravi et al., 2022; Baggen et al., 2020; Clark et al., 2010; da Silva Costa et al., 2020; Santuz et al., 2022; Sawers et al., 2017; Sawers and Bhatt, 2018) or moderate quality (Allen et al., 2019; Allen and Franz, 2018; Collimore et al., 2021; Guo et al., 2022; Hanawa et al., 2017; Kubota et al., 2021; Toda et al., 2016; Yang et al., 2019), and, two were of low methodological quality (An et al., 2013; Yang et al., 2017). The score for reporting elements was high, while external validity elements were rated lower in the studies. Four studies out of seventeen (Alizadehsaravi et al., 2022; da Silva Costa et al., 2020; Sawers et al., 2017; Sawers and Bhatt, 2018) detailed the source of patient populations.

**Table 1.**
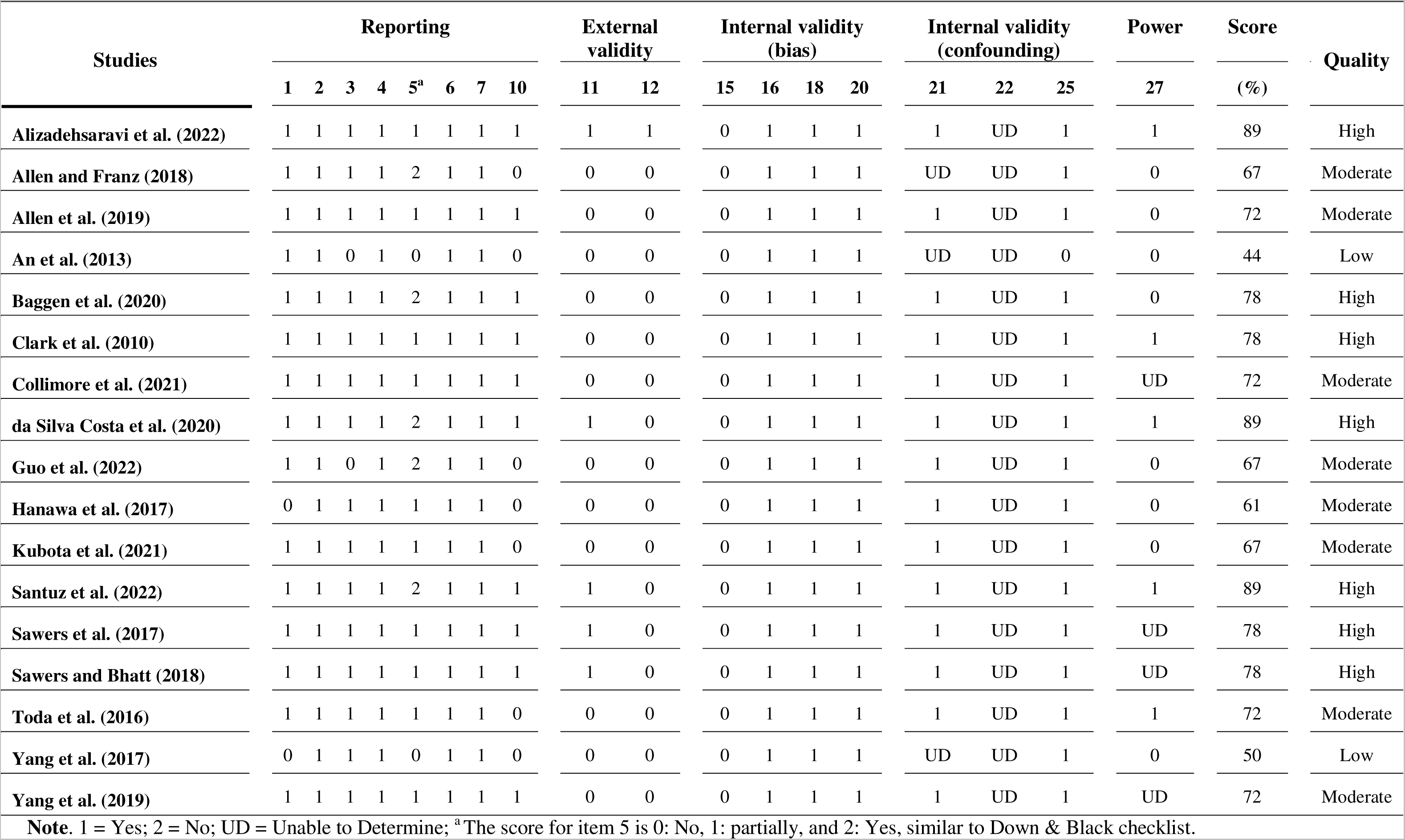
Methodological quality assessment scores of included studies using the modified version of Downs and Black checklist.

### 3.3. Studies characteristics

**Table 2** shows the population and methodology characteristics of the 17 studies included in this review. All studies were published between 2009–2022. Eleven of them (65%) used an observational cross-sectional study design (Allen and Franz, 2018; An et al., 2013; Baggen et al., 2020; Collimore et al., 2021; Da Silva Costa et al., 2020; Guo et al., 2022; Hanawa et al., 2017; Kubota et al., 2021; Santuz et al., 2022; Toda et al., 2016; Yang et al., 2017), and six studies (35%) focused on EP only (Alizadehsaravi et al., 2022; Allen et al., 2019; Clark et al., 2010; Sawers et al., 2017; Sawers and Bhatt, 2018; Yang et al., 2019). A total of 295 EP and 182 YA were included. The sample size ranged from 3 to 140 participants (group mean ± SD = EP: 17.2 ± 15.6; YA: 16.6 ± 18.6), and group age mean was 70.4 and 25.4 years old for the EP and YA adults, respectively). Eleven studies (65%) focused on muscle synergies during walking (Alizadehsaravi et al., 2022; Allen et al., 2019; Allen and Franz, 2018; Clark et al., 2010; Collimore et al., 2021; Guo et al., 2022; Kubota et al., 2021; Santuz et al., 2022; Sawers et al., 2017; Sawers and Bhatt, 2018; Toda et al., 2016), four studies (23%) on sit-to-stand task (An et al., 2013; Hanawa et al., 2017; Yang et al., 2019, 2017), one (6%) on balanced walking (Da Silva Costa et al., 2020) and one (6%) on stair ascent (Baggen et al., 2020). All studies were recording muscles using surface EMG. Overall, 5 to 16 muscles were included per leg, or leg and trunk. Seven studies (An et al., 2013; Baggen et al., 2020; Da Silva Costa et al., 2020; Guo et al., 2022; Kubota et al., 2021; Yang et al., 2019, 2017) on seventeen (41%) focused either on leg and trunk muscles. The reported muscles for the leg muscle activities were: adductor magnus (ADD), biceps femoris (BF), biceps femoris long head (BFL), biceps femoris short head (BFS), gastrocnemius (GAS), gastrocnemius lateralis (GL), gastrocnemius medialis (GM), gluteus maximus (GMax), gluteus medius (GMed), gluteus minimus (GMin), hamstrings (H), iliopsoas (IL), medial hamstrings (MH), peroneus longus (PL), rectus abdominis (RA), rectus femoris (RF), semitendinosus (ST), soleus (SOL), tensor fasciae latae (TFL), tibialis anterior (TA), vastus muscles (VAS), vastus lateralis (VL), and vastus medialis (VM). Among the seventeen included studies, the most common muscles recorded for the anterior part of the leg were: TA (n = 17; 100%), RF (n = 14; 82%), and VL (n = 15; 88%) (**Figure 2**). The most common muscles recorded for the posterior part of the leg were: SOL (n = 15; 88%), GM (n = 12; 71%), GMax (n = 11; 65%), and GMed (n = 11; 65%). The reported muscles for the trunk muscle activities were: erector spinae (ES), external obliques (EOB), latissimus dorsi (LD), paravertebral muscle (PVM), and rectus abdominis (RA). The raw EMG data was most commonly processed using the following steps: high-pass filtered, rectified, low-pass filtered, amplitude scaled, and time-normalized (see **Table 2** for more details). The majority of studies (n = 11; 65%) normalized the EMG envelopes by the maximum value (Alizadehsaravi et al., 2022; Allen et al., 2019; Allen and Franz, 2018; Baggen et al., 2020; Clark et al., 2010; da Silva Costa et al., 2020; Kubota et al., 2021; Santuz et al., 2022; Sawers et al., 2017; Sawers and Bhatt, 2018; Yang et al., 2019). However, few studies (n = 4; 23%) did not report any data normalization to extract muscles synergies (An et al., 2013; Guo et al., 2022; Toda et al., 2016; Yang et al., 2017).

**Figure 2.**
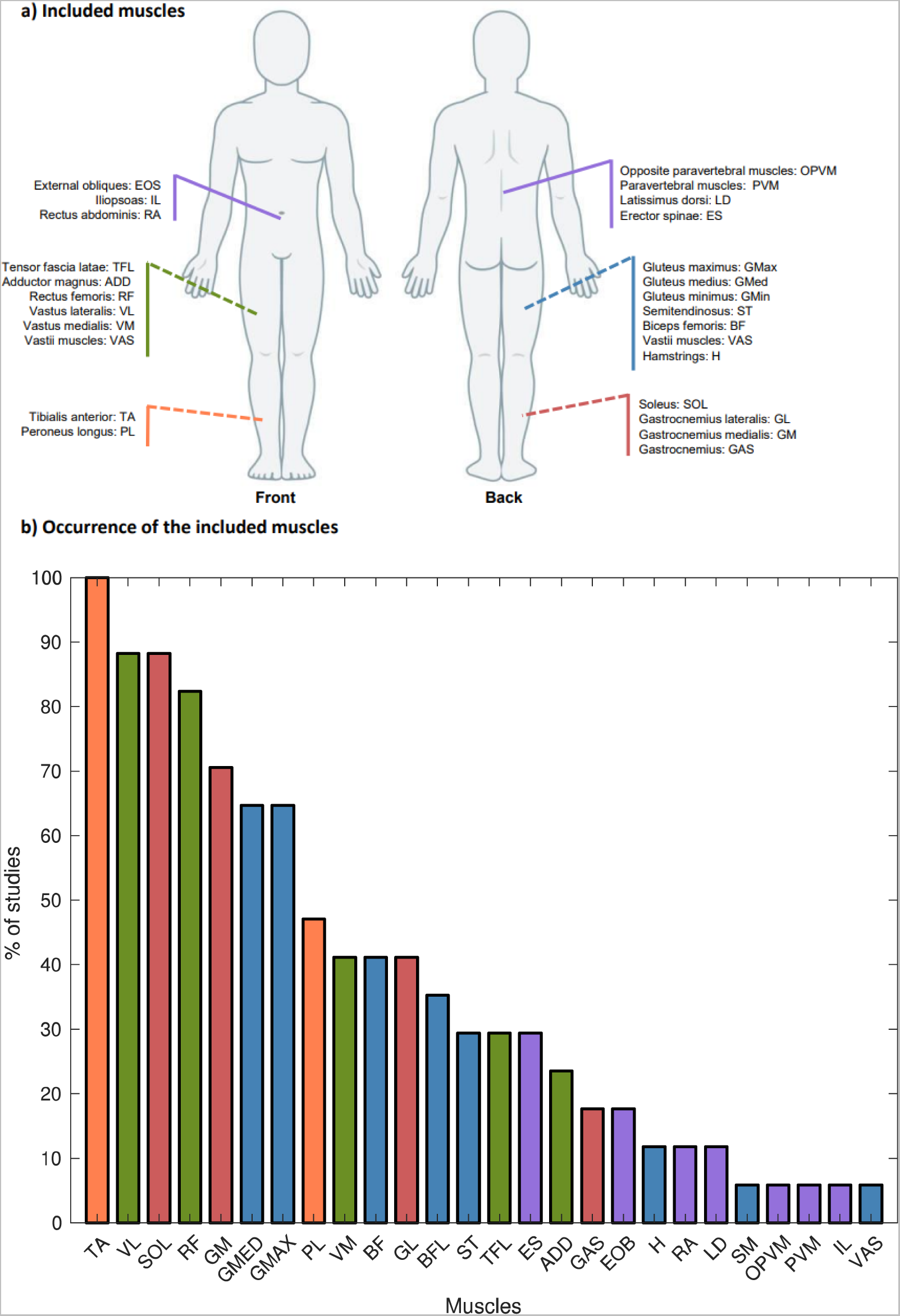
Names of the included muscles and related occurrence (% of the included studies).

**Table 2.**
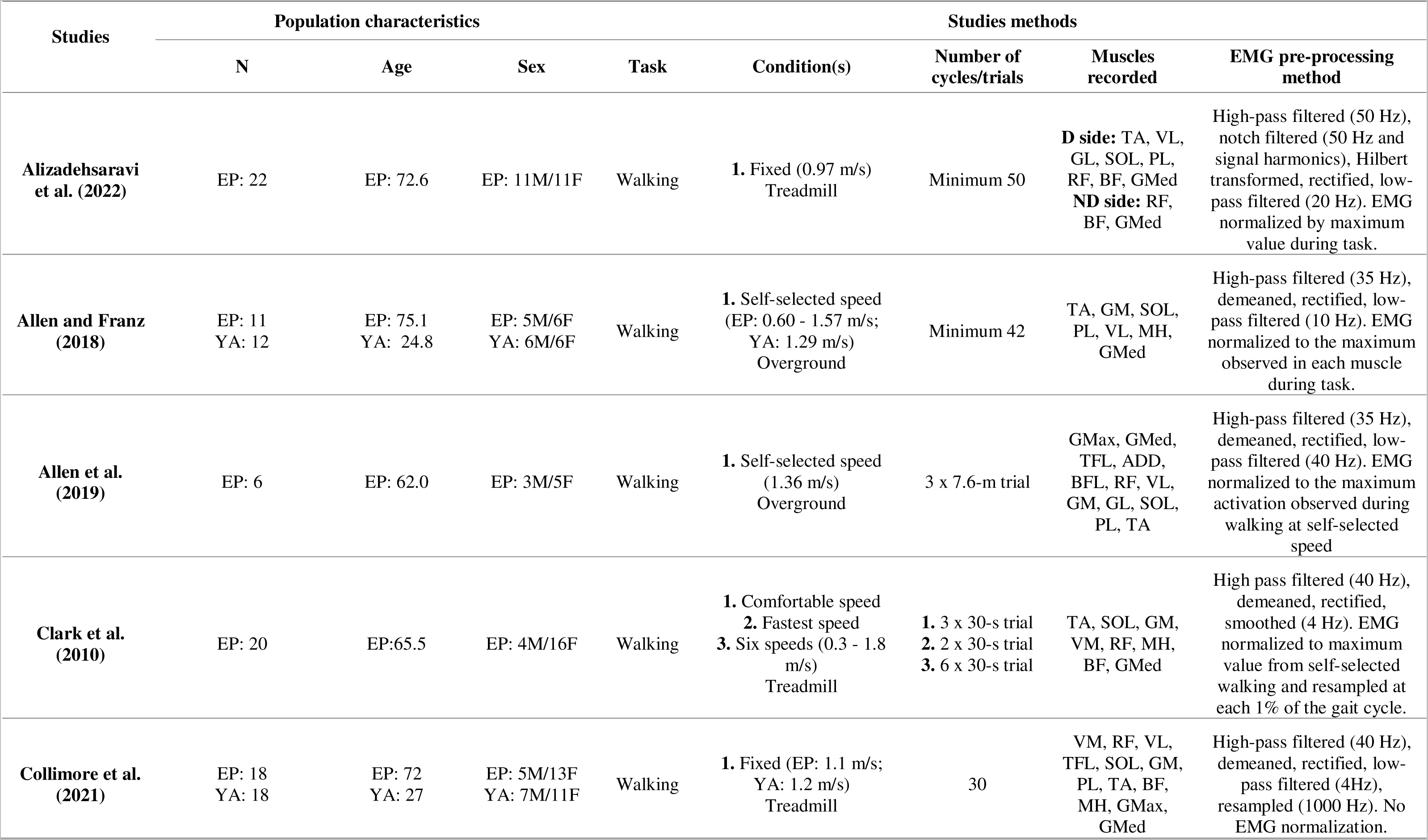
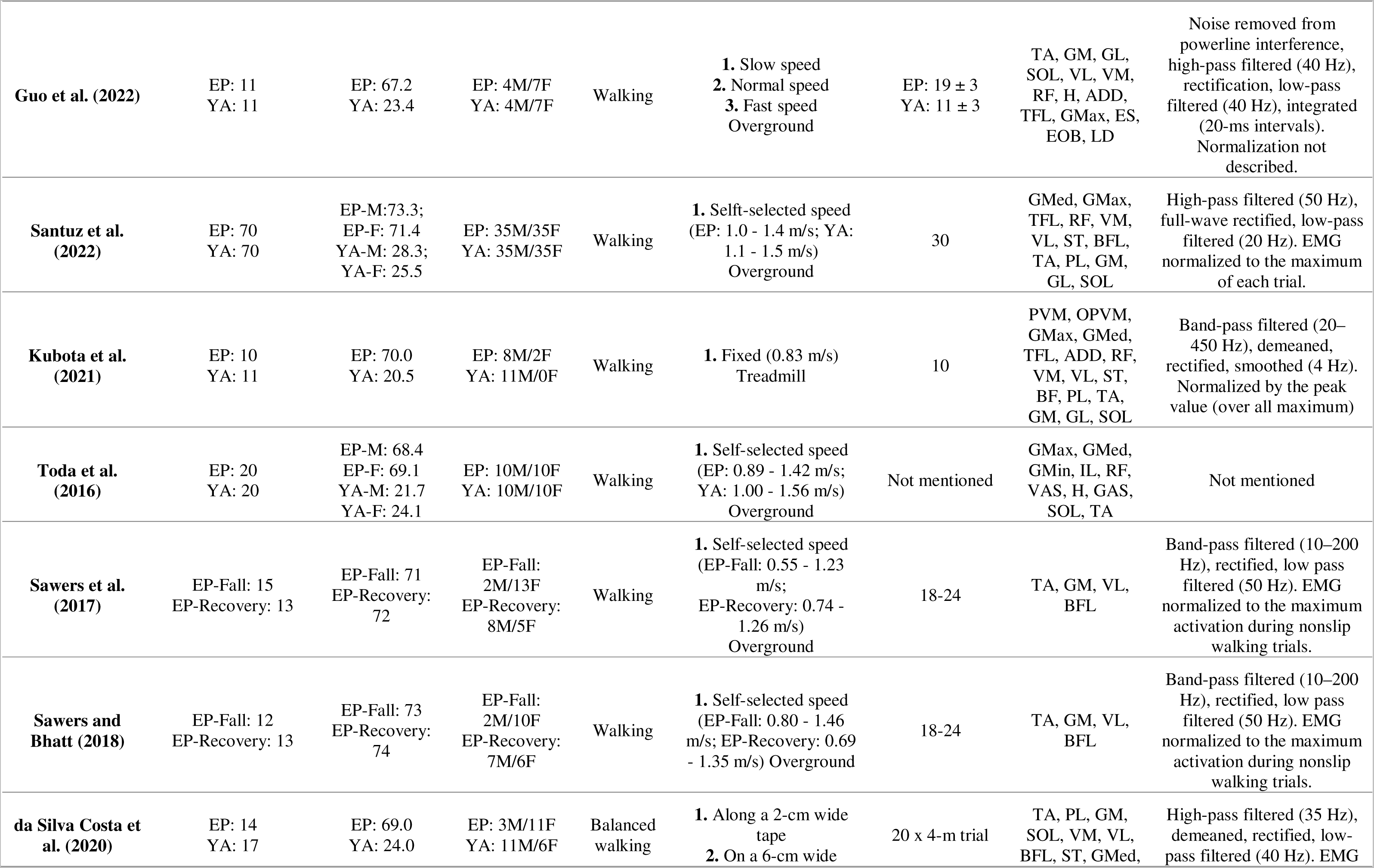
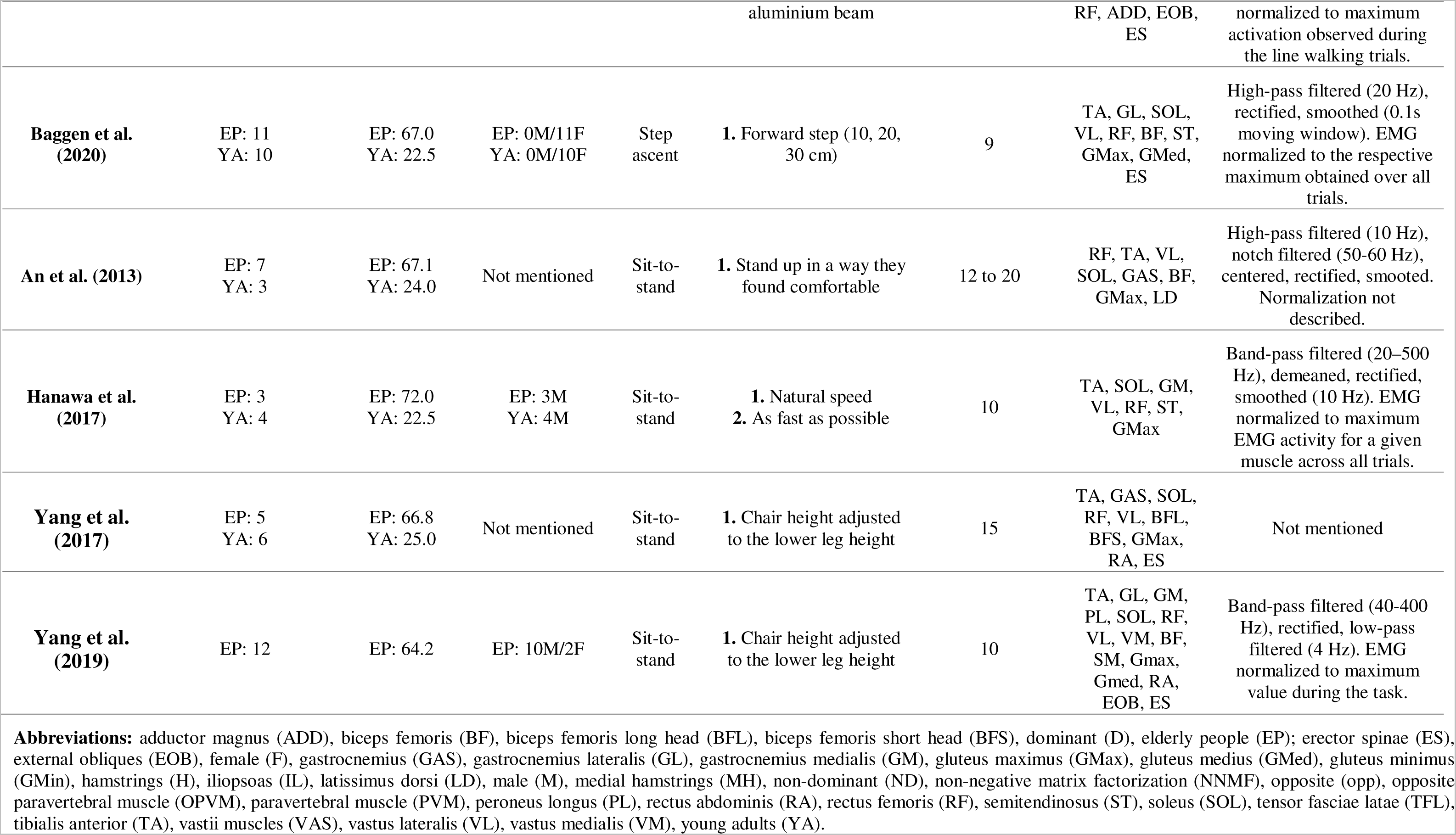
Summary of the included studies

| Studies | Population characteristics |  |  |  |  | Studies methods |  |  |
| --- | --- | --- | --- | --- | --- | --- | --- | --- |
|  | N | Age | Sex | Task | Condition(s) | Number of cycles/trials | Muscles recorded | EMG pre-processing method |
| **Alizadehsaravi et al. (2022)** | EP: 22 | EP: 72.6 | EP: 11M/11F | Walking | **1.** Fixed (0.97 m/s)<br>Treadmill | Minimum 50 | **D side:** TA, VL, GL, SOL, PL, RF, BF, GMed<br>**ND side:** RF, BF, GMed | High-pass filtered (50 Hz), notch filtered (50 Hz and signal harmonics), Hilbert transformed, rectified, low-pass filtered (20 Hz). EMG normalized by maximum value during task. |
| **Allen and Franz (2018)** | EP: 11<br>YA: 12 | EP: 75.1<br>YA: 24.8 | EP: 5M/6F<br>YA: 6M/6F | Walking | **1.** Self-selected speed (EP: 0.60 - 1.57 m/s; YA: 1.29 m/s)<br>Overground | Minimum 42 | TA, GM, SOL, PL, VL, MH, GMed | High-pass filtered (35 Hz), demeaned, rectified, low-pass filtered (10 Hz). EMG normalized to the maximum observed in each muscle during task. |
| **Allen et al. (2019)** | EP: 6 | EP: 62.0 | EP: 3M/5F | Walking | **1.** Self-selected speed (1.36 m/s)<br>Overground | 3 x 7.6-m trial | GMax, GMed, TFL, ADD, BFL, RF, VL, GM, GL, SOL, PL, TA | High-pass filtered (35 Hz), demeaned, rectified, low-pass filtered (40 Hz). EMG normalized to the maximum activation observed during walking at self-selected speed |
| **Clark et al. (2010)** | EP: 20 | EP: 65.5 | EP: 4M/16F | Walking | **1.** Comfortable speed<br>**2.** Fastest speed<br>**3.** Six speeds (0.3 - 1.8 m/s)<br>Treadmill | **1.** 3 x 30-s trial<br>**2.** 2 x 30-s trial<br>**3.** 6 x 30-s trial | TA, SOL, GM, VM, RF, MH, BF, GMed | High pass filtered (40 Hz), demeaned, rectified, smoothed (4 Hz). EMG normalized to maximum value from self-selected walking and resampled at each 1% of the gait cycle. |
| **Collimore et al. (2021)** | EP: 18<br>YA: 18 | EP: 72<br>YA: 27 | EP: 5M/13F<br>YA: 7M/11F | Walking | **1.** Fixed (EP: 1.1 m/s; YA: 1.2 m/s)<br>Treadmill | 30 | VM, RF, VL, TFL, SOL, GM, PL, TA, BF, MH, GMax, GMed | High-pass filtered (40 Hz), demeaned, rectified, low-pass filtered (4Hz), resampled (1000 Hz). No EMG normalization. |
| **Guo et al. (2022)** | EP: 11<br>YA: 11 | EP: 67.2<br>YA: 23.4 | EP: 4M/7F<br>YA: 4M/7F | Walking | 1. Slow speed<br>2. Normal speed<br>3. Fast speed<br>Overground | EP: 19 ± 3<br>YA: 11 ± 3 | TA, GM, GL,<br>SOL, VL, VM,<br>RF, H, ADD,<br>TFL, GMax, ES,<br>EOB, LD | Noise removed from<br>powerline interference,<br>high-pass filtered (40 Hz),<br>rectification, low-pass<br>filtered (40 Hz), integrated<br>(20-ms intervals).<br>Normalization not<br>described. |
| **Santuz et al.**<br>**(2022)** | EP: 70<br>YA: 70 | EP-M:73.3;<br>EP-F: 71.4<br>YA-M: 28.3;<br>YA-F: 25.5 | EP: 35M/35F<br>YA: 35M/35F | Walking | 1. Self-selected speed<br>(EP: 1.0 - 1.4 m/s; YA:<br>1.1 - 1.5 m/s)<br>Overground | 30 | GMed, GMax,<br>TFL, RF, VM,<br>VL, ST, BFL,<br>TA, PL, GM,<br>GL, SOL | High-pass filtered (50 Hz),<br>full-wave rectified, low-pass<br>filtered (20 Hz). EMG<br>normalized to the maximum<br>of each trial. |
| **Kubota et al.**<br>**(2021)** | EP: 10<br>YA: 11 | EP: 70.0<br>YA: 20.5 | EP: 8M/2F<br>YA: 11M/0F | Walking | 1. Fixed (0.83 m/s)<br>Treadmill | 10 | PVM, OPVM,<br>GMax, GMed,<br>TFL, ADD, RF,<br>VM, VL, ST,<br>BF, PL, TA,<br>GM, GL, SOL | Band-pass filtered (20–<br>450 Hz), demeaned,<br>rectified, smoothed (4 Hz).<br>Normalized by the peak<br>value (over all maximum) |
| **Toda et al.**<br>**(2016)** | EP: 20<br>YA: 20 | EP-M: 68.4<br>EP-F: 69.1<br>YA-M: 21.7<br>YA-F: 24.1 | EP: 10M/10F<br>YA: 10M/10F | Walking | 1. Self-selected speed<br>(EP: 0.89 - 1.42 m/s;<br>YA: 1.00 - 1.56 m/s)<br>Overground | Not mentioned | GMax, GMed,<br>GMin, IL, RF,<br>VAS, H, GAS,<br>SOL, TA | Not mentioned |
| **Sawers et al.**<br>**(2017)** | EP-Fall: 15<br>EP-Recovery: 13 | EP-Fall: 71<br>EP-Recovery: 72 | EP-Fall: 2M/13F<br>EP-Recovery: 8M/5F | Walking | 1. Self-selected speed<br>(EP-Fall: 0.55 - 1.23<br>m/s;<br>EP-Recovery: 0.74 -<br>1.26 m/s)<br>Overground | 18-24 | TA, GM, VL,<br>BFL | Band-pass filtered (10–200<br>Hz), rectified, low pass<br>filtered (50 Hz). EMG<br>normalized to the maximum<br>activation during nonslip<br>walking trials. |
| **Sawers and**<br>**Bhatt (2018)** | EP-Fall: 12<br>EP-Recovery: 13 | EP-Fall: 73<br>EP-Recovery: 74 | EP-Fall: 2M/10F<br>EP-Recovery: 7M/6F | Walking | 1. Self-selected speed<br>(EP-Fall: 0.80 - 1.46<br>m/s; EP-Recovery: 0.69<br>- 1.35 m/s) Overground | 18-24 | TA, GM, VL,<br>BFL | Band-pass filtered (10–200<br>Hz), rectified, low pass<br>filtered (50 Hz). EMG<br>normalized to the maximum<br>activation during nonslip<br>walking trials. |
| **da Silva Costa et**<br>**al. (2020)** | EP: 14<br>YA: 17 | EP: 69.0<br>YA: 24.0 | EP: 3M/11F<br>YA: 11M/6F | Balanced<br>walking | 1. Along a 2-cm wide<br>tape<br>2. On a 6-cm wide | 20 x 4-m trial | TA, PL, GM,<br>SOL, VM, VL,<br>BFL, ST, GMed, | High-pass filtered (35 Hz),<br>demeaned, rectified, low-pass filtered (40 Hz). EMG |
|  |  |  |  |  | aluminium beam |  | RF, ADD, EOB, ES | normalized to maximum activation observed during the line walking trials. |
| **Baggen et al. (2020)** | EP: 11<br>YA: 10 | EP: 67.0<br>YA: 22.5 | EP: 0M/11F<br>YA: 0M/10F | Step ascent | **1.** Forward step (10, 20, 30 cm) | 9 | TA, GL, SOL, VL, RF, BF, ST, GMax, GMed, ES | High-pass filtered (20 Hz), rectified, smoothed (0.1s moving window). EMG normalized to the respective maximum obtained over all trials. |
| **An et al. (2013)** | EP: 7<br>YA: 3 | EP: 67.1<br>YA: 24.0 | Not mentioned | Sit-to-stand | **1.** Stand up in a way they found comfortable | 12 to 20 | RF, TA, VL, SOL, GAS, BF, GMax, LD | High-pass filtered (10 Hz), notch filtered (50-60 Hz), centered, rectified, smoothed. Normalization not described. |
| **Hanawa et al. (2017)** | EP: 3<br>YA: 4 | EP: 72.0<br>YA: 22.5 | EP: 3M<br>YA: 4M | Sit-to-stand | **1.** Natural speed<br>**2.** As fast as possible | 10 | TA, SOL, GM, VL, RF, ST, GMax | Band-pass filtered (20–500 Hz), demeaned, rectified, smoothed (10 Hz). EMG normalized to maximum EMG activity for a given muscle across all trials. |
| **Yang et al. (2017)** | EP: 5<br>YA: 6 | EP: 66.8<br>YA: 25.0 | Not mentioned | Sit-to-stand | **1.** Chair height adjusted to the lower leg height | 15 | TA, GAS, SOL, RF, VL, BFL, BFS, GMax, RA, ES | Not mentioned |
| **Yang et al. (2019)** | EP: 12 | EP: 64.2 | EP: 10M/2F | Sit-to-stand | **1.** Chair height adjusted to the lower leg height | 10 | TA, GL, GM, PL, SOL, RF, VL, VM, BF, SM, Gmax, Gmed, RA, EOB, ES | Band-pass filtered (40-400 Hz), rectified, low-pass filtered (4 Hz). EMG normalized to maximum value during the task. |

**Abbreviations:** adductor magnus (ADD), biceps femoris (BF), biceps femoris long head (BFL), biceps femoris short head (BFS), dominant (D), elderly people (EP); erector spinae (ES), external obliques (EOB), female (F), gastrocnemius (GAS), gastrocnemius lateralis (GL), gastrocnemius medialis (GM), gluteus maximus (GMax), gluteus medius (GMed), gluteus minimus (GMin), hamstrings (H), iliopsoas (IL), latissimus dorsi (LD), male (M), medial hamstrings (MH), non-dominant (ND), non-negative matrix factorization (NNMF), opposite (opp), opposite paravertebral muscle (OPVM), paravertebral muscle (PVM), peroneus longus (PL), rectus abdominis (RA), rectus femoris (RF), semitendinosus (ST), soleus (SOL), tensor fasciae latae (TFL), tibialis anterior (TA), vastii muscles (VAS), vastus lateralis (VL), vastus medialis (VM), young adults (YA).

### 3.4. Walking tasks

A range of 4 to 8 synergies that account for more than 80% of the variance have been reported by eleven studies that have focused on <u>normal walking task</u> (see **Table 3**). Although the majority of studies (64%) (Alizadehsaravi et al., 2022; Allen et al., 2019; Allen and Franz, 2018; Clark et al., 2010; Kubota et al., 2021; Santuz et al., 2022; Toda et al., 2016) reported 4 to 5 synergies during overground or treadmill walking at different speed (speed range [max, min]: [0.30 m/s, 1.57 m/s]), one study (Collimore et al., 2021) reported only 3 synergies during treadmill walking at monitored speed (speed: 1.1 m/s), two studies (Sawers et al., 2017; Sawers and Bhatt, 2018) mentioned the presence of 6 synergies during overground walking (speed range [max, min]: [0.55 m/s, 1.26 m/s]), and one study (Guo et al., 2022) extracted 8 synergies during overground walking at different self-selected speeds (i.e., slow, normal, fast). Four of the eleven studies carried out their experiment on a treadmill, either imposing a walking speed (Alizadehsaravi et al., 2022; Collimore et al., 2021; Kubota et al., 2021), or at a self-selected speed and imposed speed (Clark et al., 2010), while seven of them conducted their experiment overground, at a self-selected (Allen et al., 2019; Allen and Franz, 2018; Guo et al., 2022; Santuz et al., 2022; Sawers et al., 2017; Sawers and Bhatt, 2018; Toda et al., 2016). All studies assessed at least the activity of 1 muscle from each sagittal lower limb muscle groups (i.e., hip flexor/extensor, knee flexor/extensor, and ankle plantar flexor/dorsiflexor), and one study (Guo et al., 2022) supplemented this with the assessment of trunk flexor/extensor, and another (Kubota et al., 2021) with the measurement of paravertebral muscles activity.

**Table 3.**
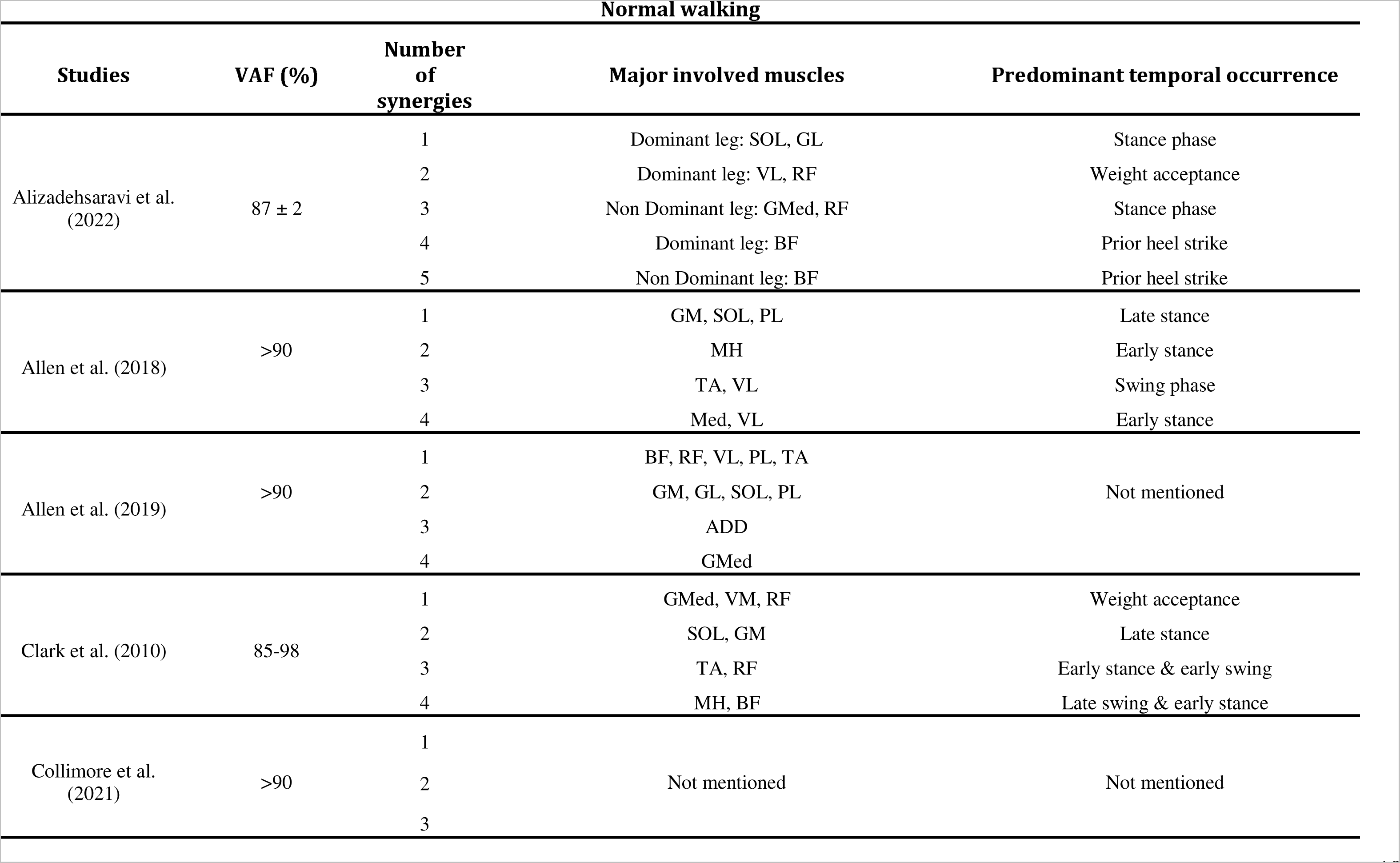
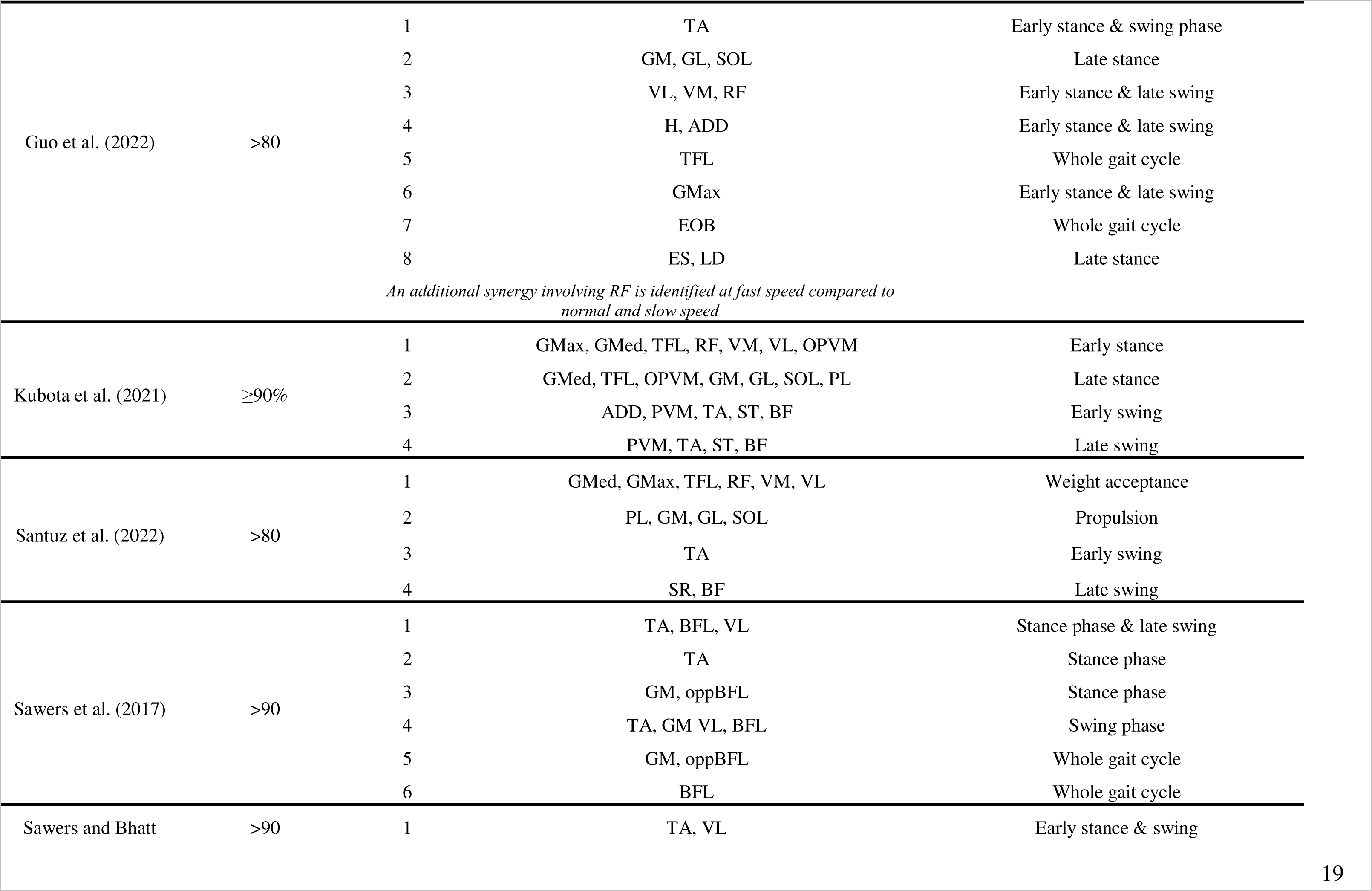
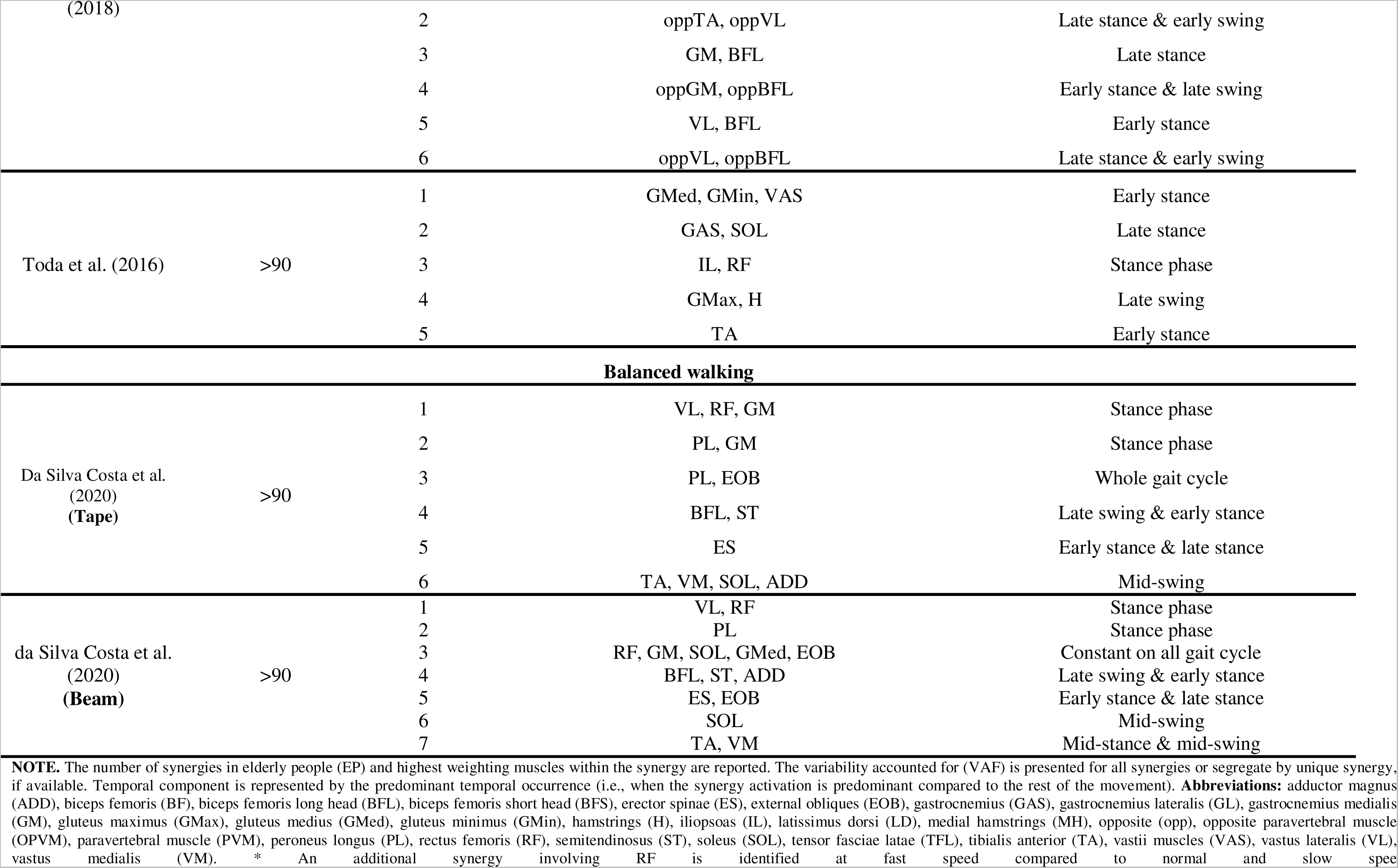
Muscle synergies during normal and balanced walking in elderly people

When comparing EP vs YA using a Dynamic Motor Control (DMC) index to identify individuals with neuromuscular complexity impairment during walking, Collimore et al. (2021) observed group difference in the number of impaired individuals. Indeed, The authors reported that 11.1% of YA (18–35 years old), 38.5% of young EP (65–74 years old) and 80% of older EP (75+ years old) presented impaired neuromuscular control (Collimore et al., 2021). Allen and Franz (2018) found that fall history, but not age, was associated with reduced number of synergies (difference: 0.90 synergy, *d* = 1.630), and greater VAF-1 (difference: +19.58 %, *d* = 2.097) (see **Table 4**). In opposition, Kubota et al. (2021) reported a reduced number of synergies for the EP group (difference: -0.87 synergy, *d* = 1.774). However, Allen and Franz (2018) highlighted that age was related to a greater synergy timing variability independent of falling history (EP-fallers difference: +1.21, *d* = 1.046, and EP-non fallers difference: +1.32, *d* = 1.396), which is in line with the results of Guo et al. (2022), who reported greater inter and intra-subject timing activation variability for most synergies at normal and slow speed. Overall, EP and YA synergies appear to differ temporally, as shown by the greater duration of activation reported by Santuz et al. (2022) for all four extracted synergies (β = 7.510 to 12.390), and the earlier shift in the activity timing of 3 of the 4 extracted synergies (β = -7.240 to 17.140). Also, our findings indicate a tendency towards a greater muscular contribution in EP for specific synergy when walking at normal speed. Indeed, Kubota et al. (2021) reported greater contribution of the ST (difference: +0.19, *d* = 0.732) and BF (difference: +0.30, *d* = 0.978) in the synergy 1 (i.e., synergy involved in loading response), Guo et al., (2022) observed a greater contribution of the ADD in the synergy 4 (i.e., synergy involved in early stance and late swing), and Toda et al. (2016) reported a greater contribution of the TA (difference: N/A, male: *d* = 0.526; female: *d* = 0.696) in the synergy 1 (i.e., synergy involved in early stance), the GMax (difference: N/A, male: *d* = 0.936; female: *d* = 0.564) and RF (difference: N/A, male: *d* = 0.021; female: *d* = 1.102) in the synergy 2 (i.e., synergy involved in late stance), the TA (difference: N/A, male: *d* = 0.261; female: *d* = 0.294) in the synergy 4 (i.e., synergy involved in late swing), and the GAS (difference: N/A, male: *d* = 0.936; female: *d* = 0.958) in the synergy 5 (i.e., synergy involved in early stance). Guo et al. (2022), who also compared differences between walking speeds, reported greater contribution of the TA and lower contribution of the TFL in the synergy 3 (i.e., synergy that contributes to loading response and leg stabilisation before the foot contact) only at fast speed, and greater contribution of the EOB in the synergy 9 only at slow speed.

**Table 4.** Significant differences in synergy characteristic between elderly people and young adults during normal and balanced walking

|  |  | VAF |  | Muscle contribution |  | Number of synergies |  | Timing activation |  | Other indexes |  |
| --- | --- | --- | --- | --- | --- | --- | --- | --- | --- | --- | --- |
|  |  | Difference in older | ES | Difference in older | ES | Difference in older | ES | Difference in older | ES | Difference in older | ES |
| Normal walking | Allen et al. (2018) | ↑ VAF-1 only in EP-fallers | 2.097 |  |  | ↓ only in EP-fallers | 1.630 | ↑ TAV - EP-fallers<br>↑ TAV - EP-non fallers | 1.046<br>1.396 |  |  |
|  | Collimore et al. (2021) |  |  |  |  |  |  |  |  | ↓ DMC index | 0.919<sup>Δ</sup> |
|  | Guo et al. (2022) |  |  | ↑ TA - S3 at fast speed<br>↓ TFL - S3 at fast speed<br>↓ TA - S4 at fast speed<br>↑ EOB - S4 at fast speed<br>↑ ADD - S4 at normal speed<br>↑ EOB - S9 at slow speed<br>↓ S6 amplitude in early swing at all speed<br>↑ S1 amplitude in late stance at fast speed | •<br>•<br>•<br>•<br>•<br>•<br>•<br>• |  |  | ↑ inter-subject TAV - S1, S2, S3, S4, S5, S8 at normal & slow speed<br>↑ intra-subject TAV - S6 at fast speed<br>↑ intra-subject TAV - S2, S4 at normal speed<br>↑ intra-subject TAV - S1 at slow speed<br>↓ intra-subject TAV - S7, S8 at normal & slow speed | •<br>•<br>•<br>•<br>• |  |  |
|  | Kubota et al. (2021) | ↑ VAF - S1<br>↑ VAF - S2<br>↑ VAF - S3<br>↑ VAF - S4 | 1.851<br>1.922<br>2.273<br>2.473 | ↑ ST - S1<br>↑ BF – S1 | 0.732<br>0.978 | ↓ | 1.774 |  |  |  |  |
|  | Santuz et al. (2022) |  |  |  |  |  |  | ↑ time of activation - S1<br>↑ time of activation - S2<br>↑ time of activation - S3<br>↑ time of activation - S4<br>Earlier shifting - S2<br>Earlier shifting - S3<br>Earlier shifting - S4 | 7.510<sup>δ</sup><br>6.530<sup>δ</sup><br>12.750<sup>δ</sup><br>12.390<sup>δ</sup><br>-7.240<sup>δ</sup><br>-15.480<sup>δ</sup><br>-17.140<sup>δ</sup> | ↑ reduction of local complexity - F vs. M<br>↑ reduction of global complexity - F vs. M | 0.040<sup>δ</sup><br>0.040<sup>δ</sup> |
|  | Toda et al. (2016) |  |  | ↓ SOL in F - S1<br>↑ TA - S1<br>↑ TA - S2<br>↑ GMax - S2<br>↑ RF - S2<br>↓ IL - S3<br>↑ GAS in F - S3<br>↑ TA - S4<br>↑ GAS - S5 | 1.405<br>M: 0.526;<br>F: 0.696<br>M: 1.039;<br>F: 0.878<br>M: 0.936;<br>F: 0.564<br>M: 0.021;<br>F: 1.102<br>M: 0.714;<br>F: 0.436<br>1.064<br>M: 0.261;<br>F: 0.294<br>M: 0.936;<br>F: 0.958 |  |  |  |  |  |  |
| Balanced walking | Costa et al. (2020) | ↑ VAF-1 | 0.840 |  |  |  |  |  |  | ↑ Wmus<br>↑ Wsum | 0.630<br>0.660 |

**NOTE.** Lower-limb muscle synergies difference in elderly people (EP) compared to young adults (YA) are presented. Synergy (S) numbers are based on Table 3. If available (• if not), effect size (ES) are reported as Cohen's d, or Glass's delta (Δ) or edges (δ). **Abbreviations:** increased/higher (↑), decreased/lower (↓), adductor magnus (ADD), bicep femoris (BF), dynamic motor control index (DMC), external obliques (EOB), females (F), gastrocnemius (GAS), gluteus maximus (GMax), iliopsoas L), males (M), soleus (SOL), temporal activation variability (TAV), tibialis anterior (TA), variance accounted for (VAF), VAF the first muscle synergy (VAF-1), number of significantly active muscles/muscle synergy (Wmus), sum of the contribution of active muscles within a muscle synergy (Wsum).

<u>As for balanced walking</u>, Da Silva Costa et al. (2020) investigated two complex walking tasks: tape and beam walking. Six and seven synergies, that accounted for more than 90% of the variance, have been extracted for the tape and beam walking conditions, respectively. Compared to YA, the authors reported higher muscle coactivation (i.e., number of significantly active muscles) within each muscle synergy (difference: +1.20 muscles, *d* = 0.630), greater muscle contribution (i.e., sum of the contributions of significantly active muscle) within a muscle synergy (difference: +0.50, *d* = 0.660), and greater VAF-1 (difference: +5.3 %, *d* = 0.840) in EP, regardless the condition (Da Silva Costa et al., 2020).

### 3.5. Stair ascent

The only study that focused on stair ascent reported 4 synergies (Baggen et al., 2020), that accounted for 90.5%, 89.8% and 91.8% of variance in young women, and 88.5%, 87.3% and 87.4% in older women for step heights of 10, 20 and 30 cm, respectively (see **Table 5**). The number of synergies was similar between step heights, and the muscle composition of Synergy 1 (i.e., synergy involved in the pull-up part of the movement), appeared to be the most variable across step heights.

**Table 5.**
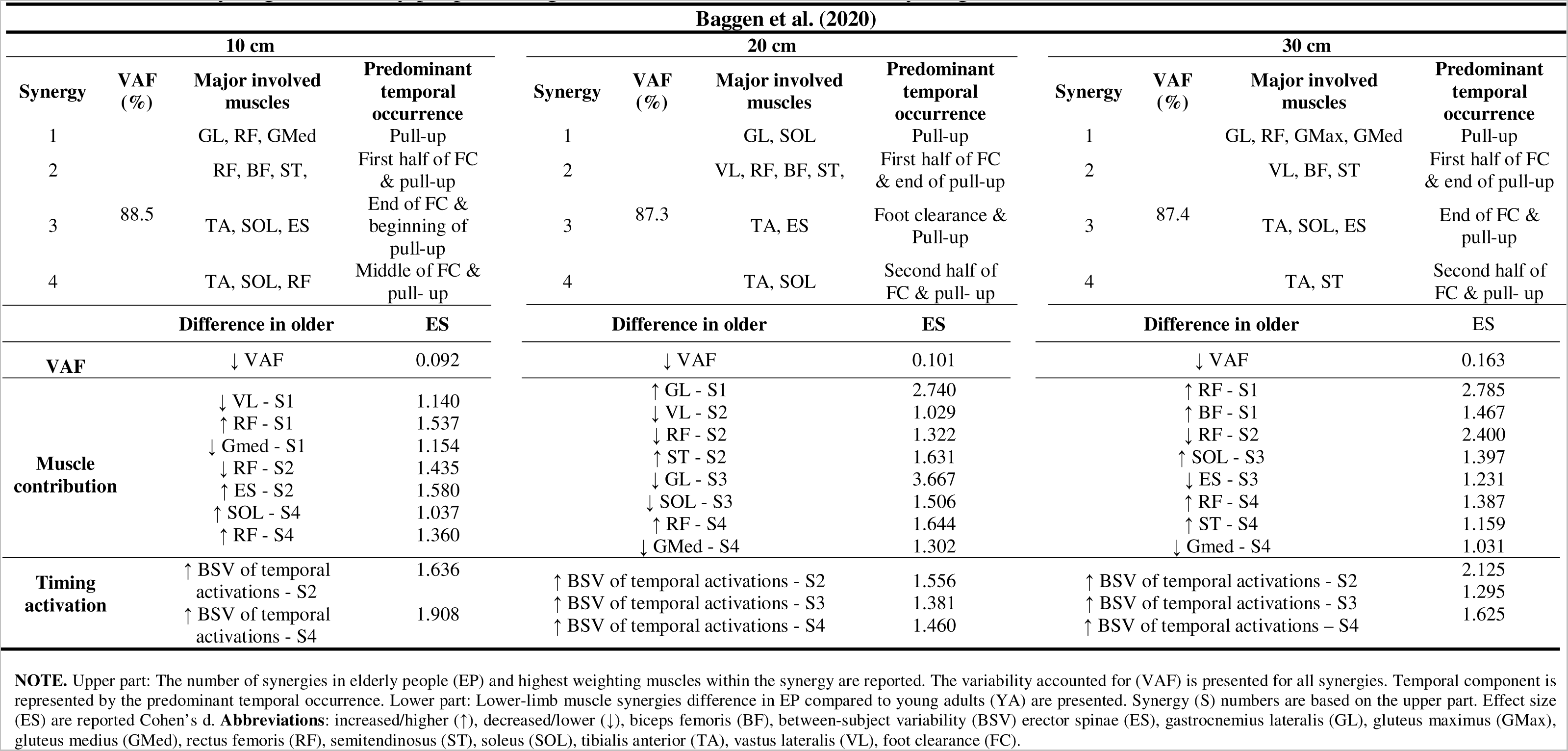
Muscle synergies in elderly people during stair ascent and differences with young adults.

The results showed that the VAF obtained when extracting 4 synergies was lower (i.e., indicating higher synergy complexity) when step height was increased, and that EP had lower VAF than YA, across all step heights (difference (10 cm): +1.92 %, *d* = 0.092; difference (20 cm): +2.53 %, *d* = 0.101; difference (30 cm): +2.88 %, *d* = 0.163). For all step heights, muscle weighting analysis showed that the RF contribution in EP is greater in the synergy 4, which is involved in the second half of foot clearance and pull-up phases (difference (10 cm): +0.21, *d* = 1.360; difference (20 cm): +0.13, *d* = 1.644; difference (30 cm): +0.20, *d* = 1.387), and lower in the synergy 2, that is contributing during the beginning of foot clearance and the end of pull-up phases (difference (10 cm): +0.26, *d* = 1.435; difference (20 cm): +0.23, *d* = 1.322; difference (30 cm): +0.36, *d* = 2.400), compared to YA. Overall, the muscle contribution differences between EP and YA appears highly variable across step heights (see **Table 5**). Regarding temporal activation patterns, higher between-subjects variability of temporal activation was shown for all step heights in the synergy 2 (difference (10 cm): +5.58, *d* = 1.636; difference (20 cm): +9.97, *d* = 1.556; difference (30 cm): +8.04, *d* = 2.125), and the synergy 4, (difference (10 cm): +6.50, *d* = 1.908; difference (20 cm): +5.36, *d* = 1.460; difference (30 cm): +9.37, *d* = 1.625) in EP, compared to YA. The same tendency was noticed, solely for 20 and 30 cm step heights, in the synergy 3, that is contributing during the end of foot clearance and the pull-up phase (difference (20 cm): +9.76, *d* = 1.381; difference (30 cm): +10.64, *d* = 1.295).

### 3.6. Sit-to-stand task

Three studies (An et al., 2013; Hanawa et al., 2017; Yang et al., 2017), among the four that focused on sit-to-stand task (An et al., 2013; Hanawa et al., 2017; Yang et al., 2019, 2017), describe the temporal occurrence of muscle synergies according to the phasic description of Schenkman et al. (1990): **Phase 1** (i.e., flexion momentum phase) begins with the first shoulder movement in the horizontal direction; **Phase 2** (i.e., momentum transfer phase) begins at contact loss with the stool; **Phase 3** (i.e., vertical extension phase) begins when the shank segment tilted forward to the maximum; **Phase 4** (i.e., stabilization phase) begins when the vertical shoulder position achieved its maximum height. A range of 3 to 4 muscle synergies that account for 88% of the variance have been reported by the four studies that have focused on sit-to-stand task (see **Table 6**).

**Table 6.** Muscle synergies in elderly people during sit-to-stand task and differences with young adults.

| Studies | VAF (%) | Number of synergies | Major involved muscles | Predominant temporal occurrence | Difference in older | ES |
| --- | --- | --- | --- | --- | --- | --- |
| **An et al. (2013)** | Not mentioned | 1 | GAS, RF, GMax | Phase 1 & 2 | 5/7 participants have no synergy at all for flexing their ankle and bending trunk - S1 | • |
|  |  | 2 | TA, VL, BF | Phase 3 |  |  |
|  |  | 3 | SOL, GAS, BF, GMax | Phase 3 & 4 |  |  |
| **Hanawa et al. (2017)** | >90 | 1 | TA, RF, VL | Phase 2 | *No information* |  |
|  |  | 2 | VL, RF, ST, GM | Phase 3 |  |  |
|  |  | 3 | SOL, GM | Phase 4 |  |  |
| **Yang et al. (2017)** | 88 ± 3 | 1 | RA | Phase 1 | *No information* |  |
|  |  | 2 | TA, VL, RF | End phase 1 & phase 2 |  |  |
|  |  | 3 | ES, VL, BFL, BFS | Phase 2 & phase 3 |  |  |
|  |  | 4 | GAS, SOL, GMax | Phase 3 & phase 4 |  |  |
| **Yang et al. (2019)** | 88.7 | 1 | RA, EOB | Phase 1 | Peak time comes after - S2<br>↓ gradient steepness after the peak value of S2 | •<br>• |
|  |  | 2 | TA, RF, VM, VL | Phase 2 |  |  |
|  |  | 3 | ES, BF, ST, GMax, GMed | Phase 3 & 4 |  |  |
|  |  | 4 | GL, GM, PL, SOL | Phase 3 and 4 |  |  |

**NOTE.** On the left side: The number of synergies in elderly people (EP) and highest weighting muscles within the synergy are reported. The variability accounted for (VAF) is presented for all synergies. Temporal component is represented by the predominant temporal occurrence. Phase 1 begins with the first shoulder movement in the horizontal direction; Phase 2 begins at contact loss with the stool; Phase 3 begins when the shank segment tilted forward to the maximum; Phase 4 begins when the vertical shoulder position achieved its maximum height (Schenkman et al., 1990). On the right side: Lower-limb muscle synergies difference in EP compared to young adults (YA) are presented. Synergy (S) numbers are based on the left side. Effect size (ES) were not available (•). **Abbreviations:** increased/higher (↑), decreased/lower (↓), biceps femoris (BF), biceps femoris long head (BFL), biceps femoris short head (BFS), external obliques (EOB), erector spinae (ES), gastrocnemii (GAS), gluteus maximus (GMax), gluteus medius (GMed), gastrocnemius medialis (GM), gastrocnemius lateralis (GL), latissimus dorsi (LD), peroneus longus (PL), rectus femoris (RF), semitendinosus (ST), soleus (SOL), tibialis anterior (TA), vastus lateralis (VL).

Overall, the results suggest similar muscle synergies underlying the sit-to-stand task between EP and YA groups. Still, An et al. (2013) observed that 5 of 7 EP had no synergy for flexing their ankle and bending their trunk (i.e., during phase 1 and 2). Also, Yang et al. (2019) reported difference in the temporal (i.e., delayed peak time), and spatial (i.e., decreased gradient steepness after peak value) structure of one synergy in EP, compared to YA. Furthermore, Hanawa et al. (2017) investigated muscle synergies during sit-to-stand task at different speeds. Three synergies, with similar spatial structure, were observed in both EP and YA groups, regardless the speed. Conversely, the change in movement speed affected the temporal structure of synergies (i.e., prolonged activation for one synergy), but no effect of age was observed.

## 4. Discussion

The goal of this scoping review was to summarize the existing literature investigating muscle synergies in EP during daily living tasks which are critical to maintaining their autonomy. We highlighted how muscle synergies in EP differ from those exhibited by YA. The main findings were: 1) EP retain in general similar muscle synergies number compared to YA although increased VAF could be observed in EP compared to YA during normal and balanced walking; 2) Generally, higher muscles contribution was reported in EP during normal and balanced walking tasks; and 3) in terms of synergies temporal structure, EP had an increased inter-subject variability during stair ascent, and an increased intra and inter-subject variability during normal walking, compared to YA.

### 4.1. Walking tasks

Despite no independent effect of age on motor module complexity in terms of number of synergies, some studies reported an age-related decrease in neuromuscular complexity (i.e., higher VAF-1) and efficiency (i.e., higher muscle coactivity) during normal walking (Allen et al., 2019; Da Silva Costa et al., 2020) and balanced walking (Da Silva Costa et al., 2020). The VAF-1 appears to be a more sensitive measure of changes in the complexity of neuromuscular control that accompanies cortical modifications related to aging (Douaud et al., 2014). Moreover, the results suggests that age increases the variability of module recruitment timing during walking (Allen et al., 2019; Guo et al., 2022), which can be associated with an altered neuromuscular control at the highest levels of control (e.g., cortical). The VAF-1 is obviously dependent on the number of synergies recruited during the task but probably also on the distribution variance and timing across the synergies from cycle-to-cycle/step-to-step. To assess these changes, that we suppose occur with age, it is likely that other synergies metrics (i.e., temporal and spatiotemporal synergy models) might provide additional information, and perhaps reveals more subtle differences. Moreover, the DMC index has been shown to be a relevant predictor related to aging, in contrast to the number of synergies (Collimore et al., 2021). Indeed, within EP group differences (young-EP vs older-EP) has been reported, suggesting that the use of two age groups (EP vs YA) as a differentiating factor can be overly crude.

Da Silva Costa et al. (2020) have shown that EP have increased muscle coactivation and contribution within synergies, which supports the increased biomechanical control demand required in EP, compared to YA. Greater antagonistic activation is a well-known adaptation when the task requires a more precise control such as walking on slippery surfaces (Chambers and Cham, 2007) or descending slopes (Lay et al., 2007), and it increases dramatically with age (Ortega and Farley, 2015). This control strategy appears intuitive, as neural delays are thought to be too long to allow feedback mechanisms to sufficiently respond to instabilities during tasks, but the effect of antagonist co-activation on control remains to be fully elucidated (Latash, 2018). Indeed, Da Silva Costa et al. (2020) have shown that task complexity, such as greater balance challenges during gait (i.e., beam vs tape walking), results in an increased muscle coactivation (i.e., within a synergy), and an increased number of muscle synergies, which reflect the greater complexity of neuromuscular activation patterns when the task is more complex. Thus, increasing the difficulty of functional task may potentially distinguished neuromuscular control deficit that are not present when the task difficulty is low (Da Silva Costa et al., 2020). Allen and Franz (2018), who assessed the effect of balanced perturbations during gait in EP with and without history of fall, suggested that fall history was an important contributor of motor module complexity. Indeed, their results indicate that fall history has a larger effect on motor module recruitment than age itself, which suggest fall experience has perceptual and biomechanical/physiological consequences on control of posture, and this likely explains the abnormal responses to balance perturbations and the associated greater risk of fall in EP with fall history observed elsewhere (Tinetti et al., 1988).

The methodological choices (i.e., environmental setting) may explain differences in EP and YA that are not supported by all studies that focussed on gait. For instance, the reduced number of synergies in EP vs. YA reported by Kubota et al. (2021) is not supported by Allen and Franz (2018) and the discrepancy could be the results of the walking environment (treadmill vs. overground), walking speed (fixed vs. self-selected), or fall history. Indeed, walking on a treadmill at a fixed speed imposes a cadenced pattern of joint motion, which may standardize the condition for the assessment of the true differences between EP and YA, despite the effect of fast speed variation that can occur during overground walking. Indeed, it could be argued that the walking environment may affects the number of synergies; Guo et al. (2022) identified an additional synergy at fast speed, compared to normal and slow speed during overground walking, while Clark et al. (2010) did not support this result with their experimentation on a treadmill. Overall, results suggest that differences in synergy outcomes are mainly observed when the conditions are the most challenging, and less observed in basic standardized conditions.

### 4.2. Stair ascent

During stair ascent task, Baggen et al., (2020) reported a decreased VAF-1 was observed in older women, and when step height was increased, suggesting a higher synergy complexity. According to the current literature, population with impaired mobility, such as cerebral palsy (Steele et al., 2015) and Parkinson’s disease (Rodriguez et al., 2013), have a reduced neuromuscular complexity, which limits their ability to perform complex locomotor task such as walking up-stairs. Thus, a greater synergy complexity in EP, compared to YA during stair ascent is not expected. However, the author proposes that the increased complexity of synergies arises from the greater challenge of stepping up stairs in EP, which requires the adoption of different control strategies to compensate for their reduced functional capacity (Baggen et al., 2020).

### 4.3. Sit-to-stand task

The ability to rise from seated position is critical for EP to maintain their independence and functional fitness (Van Lummel et al., 2015; Yee et al., 2021). In the absence of any fall history, Hanawa et al. (2017) reported common muscle synergies during sit-to-stand task in both YA (n = 4) and EP (n = 3). In the other hand, compared to YA, An et al. (2013) observed less activated synergies, and no synergy involved in ankle flexion and trunk bending (i.e., to raise the hip from the seat) in 5/7 EP. These results may be related to the low muscle strength of EP individuals, knowing that this latter is one of the most important factors to succeed in getting up from a chair (Alexander et al., 1997; Van Lummel et al., 2015). In a similar context, Van Lummel et al. (2015), showed that EP with low grip strength used a different strategy to rise from a seated position than EP with higher grip strength, which is characterized by greater trunk flexion and more dynamic trunk use during the extension phase. Indeed, peripherical muscle weakness of EP may be compensated by using their trunk, exhibiting increased sway due to the high inertia of the trunk, which is challenging to halt without control and may contribute to fall risk. However, the small sample size of studies investigating muscle synergies underlying the sit-to-stand task in EP (i.e., 3 to 12 participants) limits the results’ generalizability.

### 4.4. Clinical implications

To the best of our knowledge, this is the first review investigating the neuromuscular control as assessed by muscle synergies in EP during three common tasks: walking, sit-to-stand and stair ascent. These tasks require precise and dynamic coordination of several muscles of the lower limb and trunk. Muscle synergy analysis can provide a more generalizable assessment of motor function by identifying whether common neuromuscular control mechanisms are altered when performing multiple motor tasks. This scoping review highlighted the presence of common muscles synergies between different daily living tasks, particularly during the early stance phase of walking and the second phase of sit-to-stand task. Indeed, the concomitant activation of the TA with one or many quadriceps’ muscles (i.e., RF, VL, VM), and/or with one or many muscles of the posterior thigh (i.e., ST, BF) has been reported during early stance phase (i.e., heel strike, loading response/weight acceptance) by most of studies that focused on gait (Allen et al., 2019; Allen and Franz, 2018; Clark et al., 2010; Guo et al., 2022; Sawers et al., 2017; Sawers and Bhatt, 2018; Toda et al., 2016). Similar synergies have been observed in all studies that focused on sit-to-stand task during the second phase of the movement (Hanawa et al., 2017; Yang et al., 2019, 2017), or during the third phase (An et al., 2013). The presence of similar synergies represents a significant advantage for therapeutic managements in EP. Indeed, these results suggest the possibility to improve the neuromuscular control of problematic gait phases (e.g., during the stance phase when dynamic stability is challenged) through the practice of functional tasks that represent a lower risk of falls (e.g., chair rising with handhold).

### 4.5. Recommendations for future studies

In order that future studies may contribute to establishing a better theoretical framework concerning muscle synergies during daily living tasks in EP, authors should consider the following elements. First, the names of the muscles should be specifically defined. Some studies included in this scoping review (An et al., 2013; Guo et al., 2022; Toda et al., 2016; Yang et al., 2017) mentioned muscles group (e.g., GAS or H) without specifying which muscles were included in these groups, thereby limiting the results’ comparison with other studies. Indeed, although muscles within the same muscles group (e.g., H) work synergistically during the movement (e.g., walking), the contribution of each muscle (e.g., BF, ST, SM) may vary depending on the specific demands of the gait phase.

Second, the number and the choice of the muscles included in the analysis should be chosen carefully. Indeed, Steele et al (Steele et al., 2015) have shown that the structure of synergies is dependent of the number and the muscles choice in the analysis. They have reported that the VAF is over-estimated when fewer muscles are included in the analysis (Steele et al., 2015). In line with this latter and the results of this scoping, we recommend to 1) record the muscle activity of as many muscles as possible that are involved in the task (i.e., muscles recognized as reliable through surface EMG recording), and to 2) select the largest muscles (i.e., determined by maximum isometric force (Maughan et al., 1983)) if the number of muscles that can be recorded is limited (Steele et al., 2015).

Third, attention should be given to minimizing noise when recording EMG signals. Indeed, Steele et al. (2015) have shown that noise affects the synergy analysis outcomes when a small number of muscles are recorded. However, when the analysis includes more than 15 muscles, they reported that a signal that is at least 10 times stronger than the noise has minimal effect on muscle combinations (Steele et al., 2015). Noise can be limited, for instance, by a good skin preparation (i.e., shaving, sandpapering and cleaning of the skin), by choosing the best location and orientation of the sensor on the muscle (i.e., region away from the innervation zone and the end zone of the muscle), and by good fixation of the sensor (i.e., with elastic band or tape) (Hermens et al., 2000).

Fourth, there is no one-size-fits-all method for signal processing, but authors should be aware of the effect of the signal preprocessing on synergies estimation. For instance, Guo et al., (2022), who used an unconventional signal processing technique (i.e., a quasi-raw pattern/lightly filtered, and cluster synergies), found a total of 8 synergies, which is largely above the number of synergies conventionally reported (i.e., n = 4 to 5) by previous studies focusing on walking task (Alizadehsaravi et al., 2022b; Allen et al., 2019b; Allen and Franz, 2018b; Clark et al., 2010b; Kubota et al., 2021; Santuz et al., 2022b; Toda et al., 2016b).

Finally, the VAF-1, the number of synergies or the DMC are interesting, but too gross and indiscriminating. An analysis of the synergy’s spatial and temporal structure is warranted to detect neuromuscular control deficits. A conventional EMG analysis can provide an overview of the spinal cord’s output, and the synergy analysis allows identifying its structuring. The following three approaches, that are not well reported in the current literature, would be interesting to consider: 1) assessing neuromuscular robustness over time throughout a task, by calculating the variability using the cross-VAF (Ghislieri et al., 2023; Gizzi et al., 2015). This would make it possible to describe how an epoch containing N gait cycles can be reconstructed by a muscle synergy model calculated from a different epoch of N gait cycles. 2) Using the intra-subject approach, and 3) an inter-subject/repertory approach, which both allow a better synthetization of results across subjects or condition (Cheung et al., 2020; Funato et al., 2022; Guo et al., 2022a). The intra-subject approach is promising, but the inter-subject comparison is questionable in the absence of adequate EMG signals normalization, since synergies may differ because of the normalization technique used or the different muscle activation level.

### 4.6. Limitations

Regarding the review itself, few limitations must be acknowledged. A limitation concerns the restrictions on the language, and the type of publication. Indeed, we have chosen to include only English and French publications to ensure that we fully comprehend the content of the articles and accurately extract relevant information. Regarding the qualities of included studies, several studies included a limited number of participants, without performing prospective sample size calculations. Therefore, some of them may not have the power to detect changes in muscle synergies. Also, two of the four studies that focused on sit-to-stand task have a low methodological quality assessment score, which constrained the quality of our results concerning this latter. However, the majority of the included studies have a large sample size and a moderate to high methodological quality assessment score. Finally, the metrics used to describe synergies are numerous (e.g., VAF, VAF-1, DMC, TAV), and these methodological discrepancies limit the information synthetization across studies.

## 5. Conclusion

The identification and analysis of muscle synergies provide insights into the coordination and functional implications of muscle activation patterns during common daily activity. This approach was prioritized in this review to understand age-related changes in neuromuscular control when performing daily living tasks such as walking, sit-to-stand and stair ascent. Our findings suggested that although the number of synergies remains similar between YA and EP, other metrics such as DMC, VAF, and spatial and temporal structures of synergies enable the identification of decline in neuromuscular control in EP.

## Authors’ contributions

**YC** developed the search strategy and methodology for this review, which have been validated by a science librarian**. YC** and **SH** screened the search hits for eligibility and rated the quality of the included studies. **CDP** and **YC** extracted and synthesized the relevant data. **CDP** wrote the first draft of the manuscript. **YC**, **NT**, **KT** and **FM** performed a major revision of the manuscript. All authors have read and agreed to the published version of the manuscript.

## Data Availability

not applicable

## Acknowledgments

C.D.P. is scholar from Fonds de recherche du Québec—Santé (FRQS). The authors thank Martine Gagnon (a science librarian at Laval University) for her guidance and advice during the implementation of the research strategy.

